# Automatic Physical Examination Segmentation within Objective Structured Clinical Examination Videos

**DOI:** 10.1101/2025.04.03.25325195

**Authors:** Shinyoung Kang, Michael J. Holcomb, David Hein, Ameer H. Shakur, Thomas O. Dalton, Andrew R. Jamieson

**Author notes:** Corresponding Author: Andrew Jamieson.

## Abstract

**Objective:** Assessing medical student performance in Objective Structured Clinical Examinations (OSCEs) is labor-intensive, requiring trained evaluators to review 15-minute long videos. The ‘physical examination’ period constitutes only a small portion of these videos. Automated segmentation of OSCE videos could significantly streamline the evaluation process by detecting this physical exam portion for targeted evaluation. Current video analysis approaches struggle with these long recordings due to computational constraints and challenges in maintaining temporal context. This study tests whether multimodal large language models (MM-LLMs) can segment physical examination periods in OSCE videos without prior training, potentially easing the burden on both human graders and automated systems.

**Methods:** We analyzed 500 videos from five OSCE stations at UT Southwestern Simulation Center, each 15 minutes long, using hand-labeled physical examination periods as ground truth. MM-LLMs processed video frames at one frame per second, classifying them into discrete activity states. A hidden Markov model with Viterbi decoding ensured temporal consistency across segments, addressing the inherent challenges of frame-by-frame classification.

**Results:** Using Viterbi decoding trained on just 50 hand-labeled videos (10 from each station), zero-shot GPT-4o achieved 99.8% recall and 78.3% intersection over union (IOU), effectively capturing physical examinations with an average duration of 175 seconds from 900-second videos—an 81% reduction in frames requiring review.

**Conclusions:** Integrating multimodal large language models with temporal modeling effectively segments physical examination periods in OSCE videos without requiring extensive training data. This approach significantly reduces review time while maintaining clinical assessment integrity, demonstrating that zero-shot AI methods can be optimized for medical education’s specific requirements. The technique establishes a foundation for more efficient and scalable clinical skills assessment across diverse medical education settings.

## Objective

### Background

Objective Structured Clinical Examinations (OSCEs) have been integral to medical education assessment since their introduction in the 1970s, offering a standardized method to assess clinical competencies [1–3]. During these exams, medical students rotate through multiple 15-minute stations, interacting with standardized patients (SPs) to demonstrate skills in communication, physical examination, and clinical reasoning. This controlled setting ensures consistent evaluation across diverse scenarios, such as pediatric assessments or surgical consultations.

Grading OSCE performance relies heavily on trained standardized patient evaluators (SPEs) who review video recordings, assessing students against rubrics that emphasize verbal communication, emotional intelligence, and physical examination proficiency. This manual process is labor-intensive and resource-demanding, creating a significant bottleneck as medical schools scale training for larger cohorts.

While automated assessment has progressed for written components of medical examinations, leveraging natural language processing for transcript analysis and written exams [4–6], video-based evaluations like OSCEs remain challenging to automate [7, 8]. The complexity stems from the dynamic, visual nature of clinical interactions, including subtle movements and gestures that require deep domain understanding. Existing multimodal models struggle with 15-minute OSCE recordings due to computational constraints and challenges maintaining temporal context over extended durations, as models must either truncate sequences or aggressively downsample, sacrificing critical details [9–11]. These limitations are particularly pronounced for physical examinations, where students’ practical skills must be precisely evaluated. Physical examination assessment, therefore, remains a critical gap in automated OSCE evaluation, and identifying and isolating these segments is a foundational step toward comprehensive automated assessment of clinical competencies.

### Significance

To address this gap, we propose a novel approach that fundamentally differs from general video classification methods. Unlike traditional approaches that assign a single label to entire recordings [12, 13], our method performs temporal segmentation to isolate specific activities within OSCE videos precisely. This distinction is critical in educational contexts, where evaluators need to assess discrete skills demonstrated in brief segments, such as the physical exam portion, which typically constitutes only 15-20% of the total recording time.

Despite the critical importance of physical examination assessment in medical education, to our knowledge, this study represents the first attempt to automatically segment physical examination periods within OSCE videos. While temporal segmentation approaches have been employed in other domains [14], the unique requirements of OSCE evaluation—particularly the need for near-perfect recall to capture even brief examination components—present distinct challenges that general methods were not designed to address.

Our approach integrates multimodal large language models (MM-LLMs) with Viterbi decoding through hidden Markov models, creating a solution specifically tailored to medical education’s requirements. This combination leverages the contextual understanding capabilities of MM-LLMs for frame-level classification while applying Viterbi decoding to filter out spurious false positives and negatives, ensuring consistent segmentation of examination activities across the video timeline. Our primary research question is: Can MM-LLMs accurately identify physical examination activities in OSCE videos without specialized training across diverse examination scenarios?

We evaluate our approach across 500 recordings from five OSCE stations at UT Southwestern Simulation Center: Family Medicine, Surgery 1, Surgery 2, Internal Medicine 1, and Internal Medicine 2. Each station presents unique challenges, from differing examination techniques to varied spatial configurations. Additionally, each station utilizes different camera angles due to variations in room setup, allowing us to test the robustness of our approach across diverse recording environments.

By streamlining video analysis, our method not only enhances grading efficiency but also establishes a foundation for fully automated OSCE assessment. This advancement aligns with the broader shift toward competency-based medical education frameworks, where efficient, objective assessment of clinical skills is increasingly vital. As medical education evolves to meet growing healthcare workforce demands, such technological innovations could significantly improve the scalability and consistency of clinical skills evaluation while alleviating institutional resource constraints.

## Methods

### Study Design and Dataset

This study analyzed Objective Structured Clinical Examination (OSCE) videos from medical students at the UT Southwestern Simulation Center. We specifically focused on the Comprehensive OSCE (COSCE) administered during clerkship. Of the ten total OSCE stations administered in the fall of 2019, we selected the five that included physical examination (PE) components: Family Medicine, Surgery 1, Surgery 2, Internal Medicine 1, and Internal Medicine 2. For each station, we randomly selected 100 videos (500 videos total), each approximately 15 minutes (900 seconds) in length.

The videos were recorded at 1920×1080 resolution with two fixed cameras: one facing the patient and one facing the medical student. For our analysis, we primarily used the camera angle facing the patient to best capture the medical student’s movements during physical examination.

### Ground Truth Labeling

To establish ground truth for model evaluation, a single rater with Emergency Medical Technician level expertise manually annotated the physical examination periods in all 500 videos. We defined the physical examination period using clear visual criteria: beginning when the medical student makes direct physical contact with the patient for assessment purposes (such as when the otoscope light aligned with the patient’s eye during an eye examination) and ending when all direct physical assessment activities concluded. This definition excluded preparatory activities like retrieving examination tools, ensuring that only unambiguous examination activities were labeled. Given the straightforward nature of identifying direct physical contact beginning and ending points, a single expert annotator was sufficient for this task.

### Automated Physical Examination Segmentation Workflow

Our approach employs a streamlined pipeline for identifying physical examination periods within OSCE videos. This process consists of human detection preprocessing, direct frame classification using multimodal large language models (MM-LLMs), and temporal consistency enforcement through Viterbi decoding. The complete workflow is illustrated in Figure 1.

**Figure 1.**
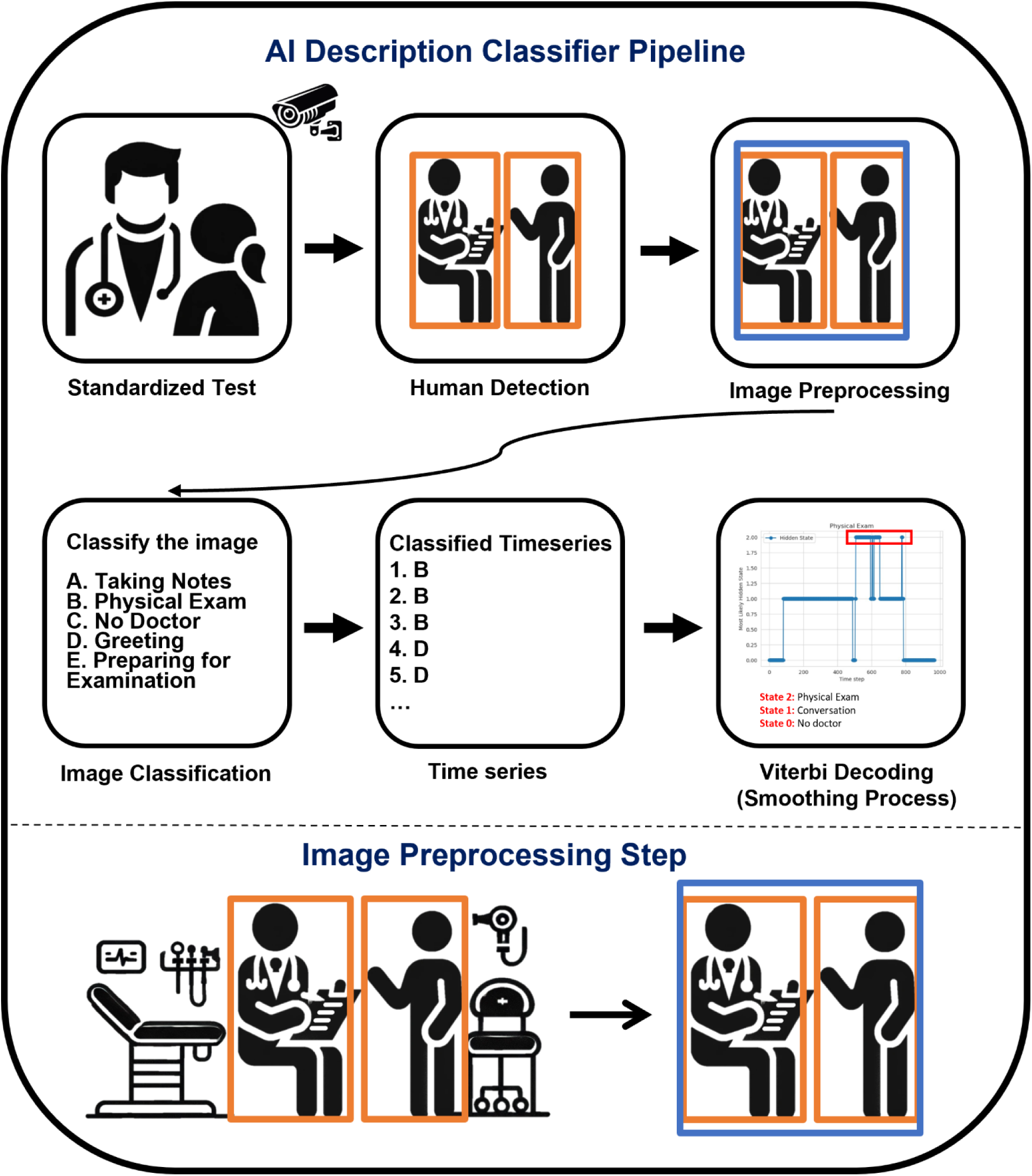
Frame Classification pipeline for physical exam detection. This streamlined approach classifies frames directly into states (Taking Notes, Physical Exam, No Doctor, Greeting, or Preparing for Examination) before applying Viterbi decoding.

### Preprocessing with Human Pose Detection

We used YOLOv11-pose to detect human presence in each frame [15, 16]. This preprocessing step allowed us to optimize computational efficiency by only having the multimodal LLM process frames containing people. Frames without detected humans were automatically assigned to the “No Doctor” state.

The model identified the bounding boxes of human skeletons and was configured to detect a maximum of two individuals per frame (the medical student and standardized patient), which aligned with the design of OSCE systems. We set a minimum confidence threshold of 0.5 for human detection, balancing detection sensitivity with computational efficiency.

For each frame containing human subjects, we created a square bounding box encompassing all detected individuals. As illustrated in Figure 1, these cropped frames were then resized to 512×512 pixels for MM-LMMs.

### Frame Classification Approach

We performed frame analysis at three different sampling rates to evaluate computational efficiency versus accuracy tradeoffs: one frame every second (1s interval), one frame every two seconds (2s interval), and one frame every three seconds (3s interval). These specific sampling rates were selected to balance computational requirements with detection accuracy. The lower bound of 3s intervals was particularly significant, as physical examinations in OSCEs can sometimes be brief (2-3 seconds for specific techniques). Sampling less frequently would risk missing these short examination components entirely.

For frame classification, we evaluated six multimodal large language models: GPT-4o-11-06, GPT-4o-mini, Gemini-2.0-Flash-exp, Gemma-3, Qwen-2.5VL-72b, and Qwen-2.5VL-7b. Open-source models were hosted on graphics processing unit nodes within BioHPC, UTSW’s shared high-performance computing infrastructure. Proprietary models were accessed via secure, HIPAA- and FERPA-compliant APIs. These models analyzed each preprocessed frame using the structured prompt depicted in Table 2.

**Table 1.** Video count and physical examination duration statistics by station.

|  | Count | Average Physical Exam Time (sec) | Standard Deviation of Physical Exam Time (sec) |
| --- | --- | --- | --- |
| Overall | 500 | 126 | 61 |
| Family Medicine | 100 | 141 | 70 |
| Surgery 2 | 100 | 163 | 74 |
| Internal Medicine 1 | 100 | 108 | 54 |
| Surgery 1 | 100 | 119 | 59 |
| Internal Medicine 2 | 100 | 101 | 47 |

**Table 2.**
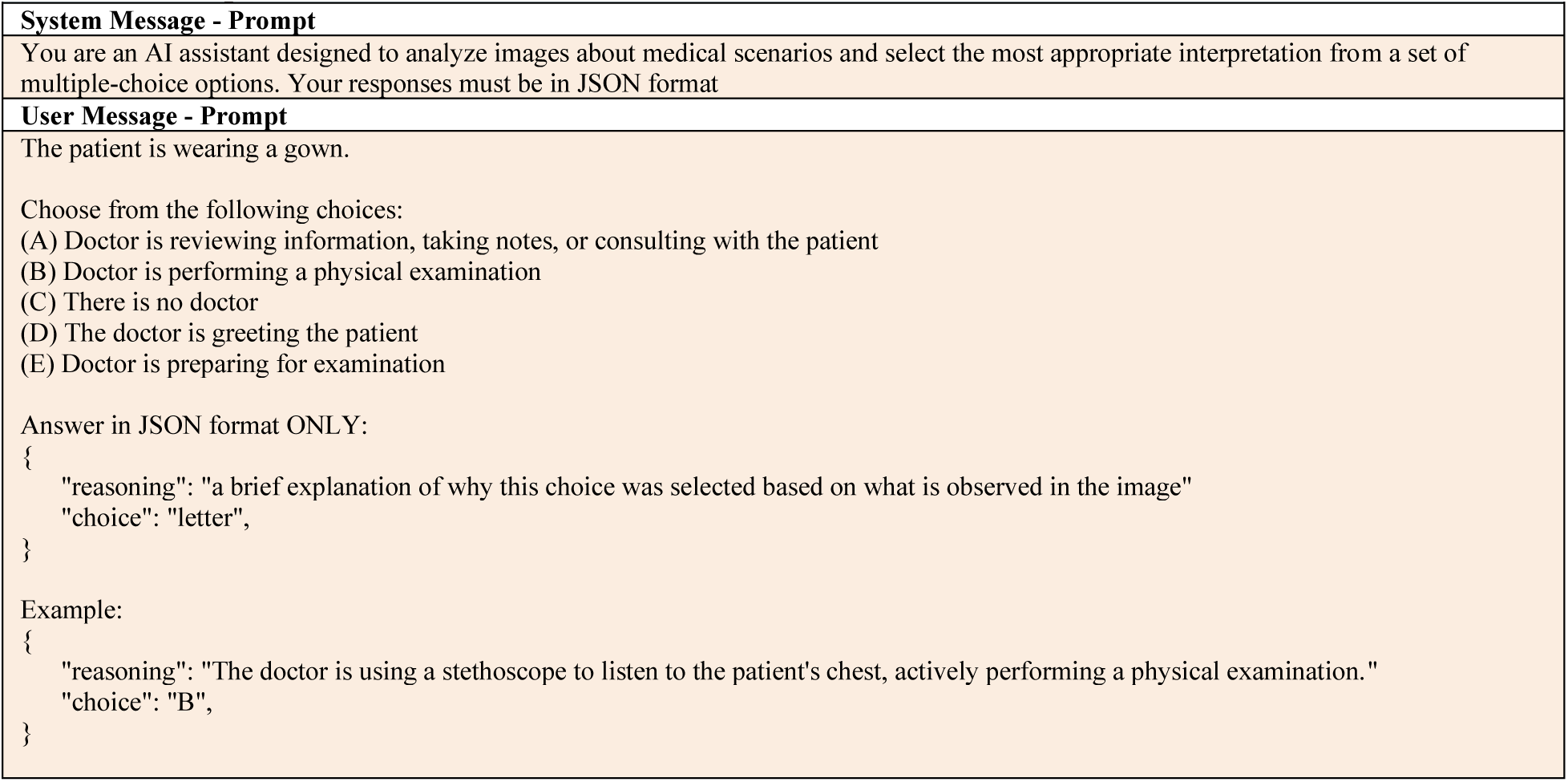
Prompts for Frame Classification Task.

### Viterbi Decoding for State Prediction

After obtaining frame-level classifications, we implemented a hidden Markov model (HMM) with Viterbi decoding to enforce temporal consistency and address the inherent noise in frame-level predictions. We simplified the analysis by merging the five initial categories into three primary states that reflect the fundamental activities in OSCE encounters (Figure 2 A): Taking Notes/Consulting, Physical Examination, and No Doctor.

**Figure 2.**
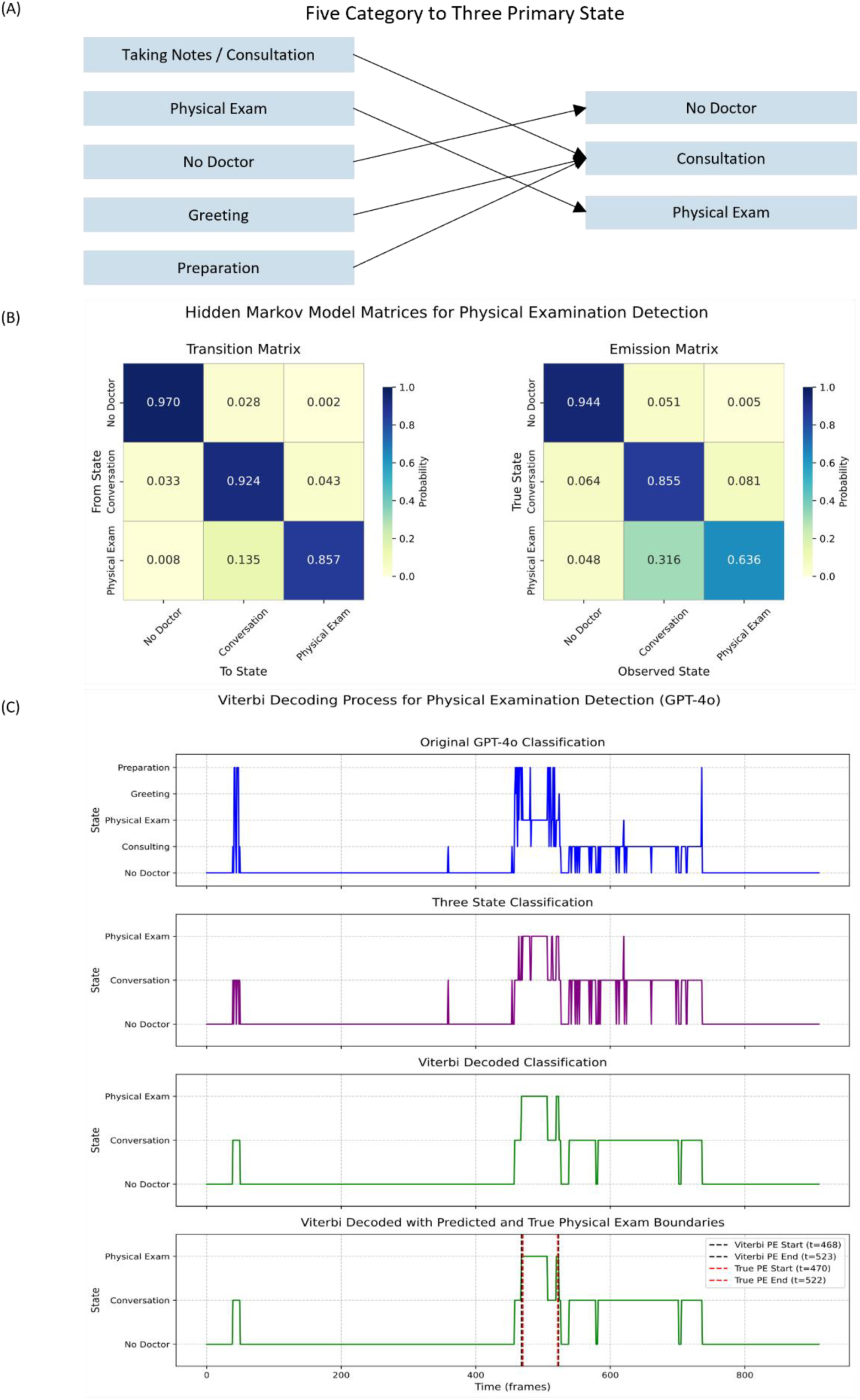
Viterbi Decoding Process. (A) Five Category to Three Primary States and (B) Hidden Markov Model Matrices: transition matrix showing probabilities between states and emission matrix showing a relationship between true and observed states. (C) Example of the Viterbi Decoding Process for Finding Physical Exam within a Video

We randomly selected 10 videos from each station (50 videos total) to calculate the average transition and emission matrices for the HMM shown in Figure 2B. The transition matrix captured the probability of moving between states, while the emission matrix represented the probability of observing a particular state given the true underlying state.

Based on the OSCE protocol, we initialized the model with “No Doctor” as the starting state since sessions begin with the patient alone in the examination room. We then applied the Viterbi algorithm through the PyHHMM library [17] to determine the most likely sequence of states and identify the physical examination period.

To ensure complete capture of physical examination periods despite discrete sampling, we added a buffer time at the edges of detected periods proportional to the frame sampling rate (Figure 3). This buffer compensated for transitions that might occur between sampled frames. Additional experiments examining the effect of different buffer times (0, 15, and 30 seconds) on performance metrics are presented in Appendix A, demonstrating how increased buffer sizes improve recall at the expense of precision and IOU.

**Figure 3.**
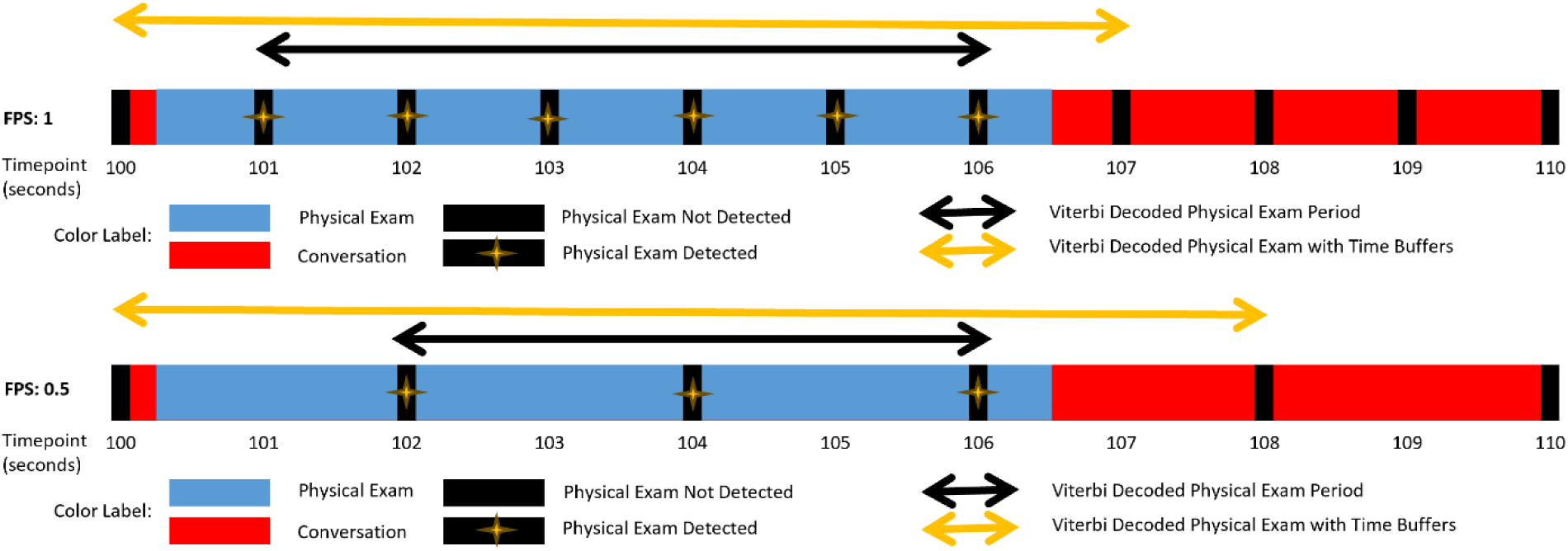
Buffer time implementation to ensure complete transition capture between sampled frames.

### Evaluation Metrics

We assessed model performance using four primary metrics, calculated per video and averaged across all videos:

1. **Recall**: The proportion of the labeled physical examination period correctly captured by the model. A high recall is critical, as missing any PE segment compromises clinical skill assessment.

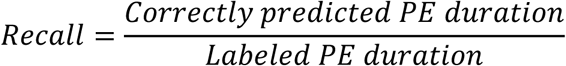 Recall =1: Complete capture of physical examination Recall =0: None captured
2. **Precision**: The accuracy of the predicted PE segments, calculated as the ratio of correctly predicted PE duration to total predicted PE duration.

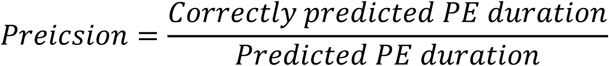
3. **Intersection over Union (IOU):** Measures the overlap accuracy between predicted and actual PE periods.

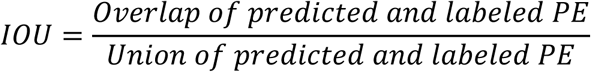
4. **Predicted PE Length:** Evaluates practical utility by quantifying the potential reduction in video review time.

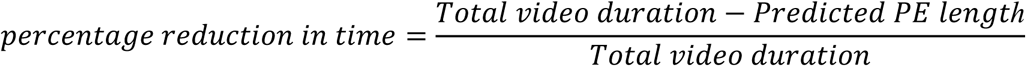

We prioritized high-recall models, accepting slightly longer predictions to ensure critical examination segments were not omitted. All metrics were calculated on a per-video basis and averaged across the dataset, with 95% confidence intervals reported.

## Results

### Overall Model Performance Across Sampling Rates

Our evaluation compared hand-labeled physical examination (PE) periods against those identified through our approach across five OSCE stations (n=500 videos). We prioritized recall as our primary metric, as missing examination components could lead to incomplete evaluation of clinical skills in high-stakes educational assessments. Table 3 presents comprehensive findings across all models and sampling rates.

**Table 3.**
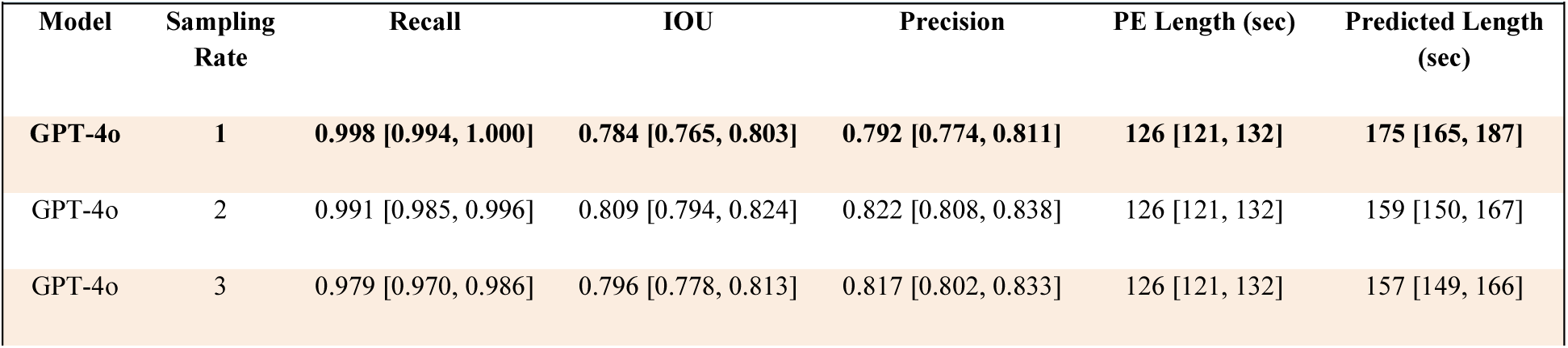
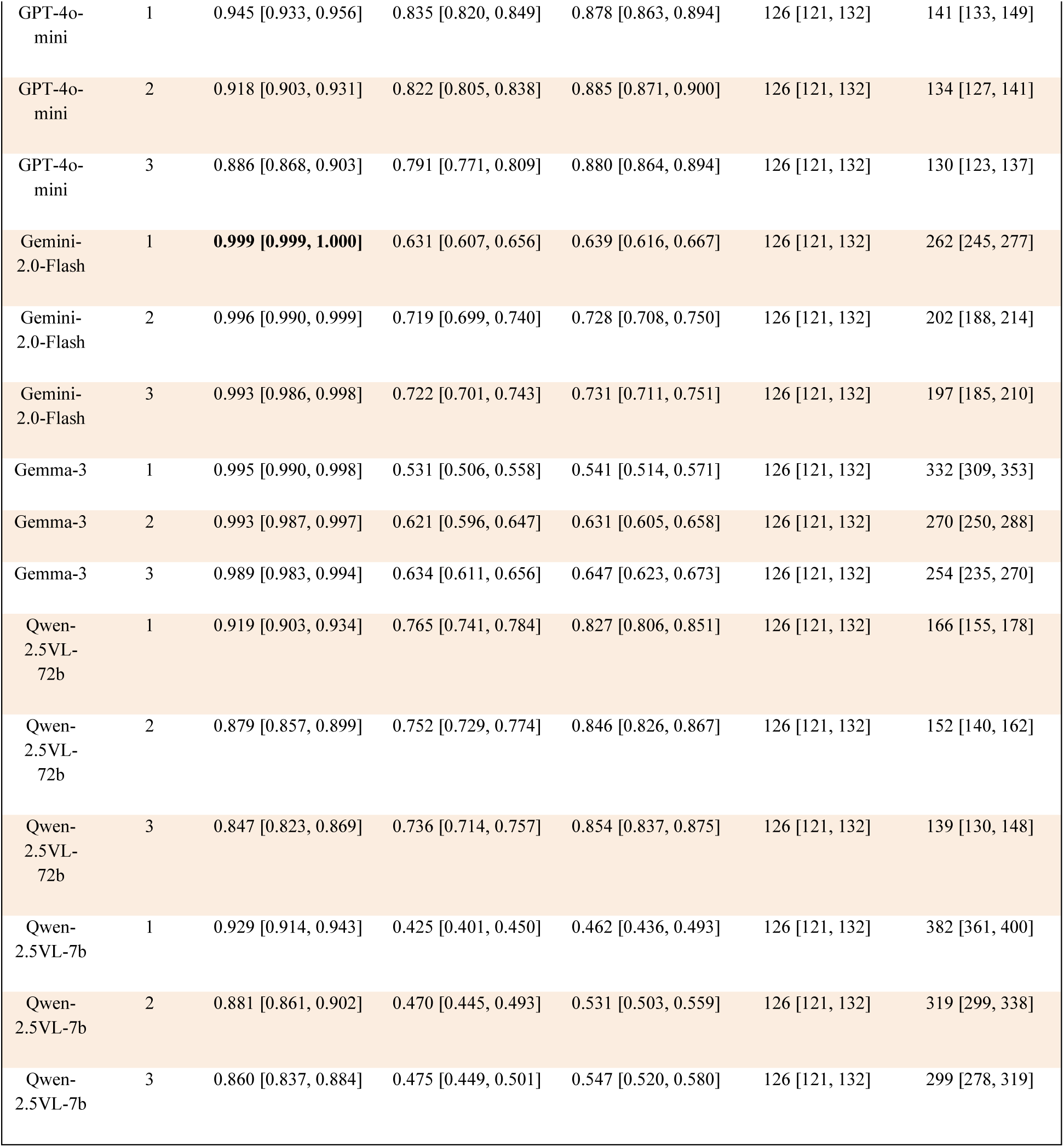
Overall Performance Comparison by Sampling Rate with 95% CI.

| Table 3. Overall Performance Comparison by Sampling Rate with 95% CI |  |  |  |  |  |  |
| --- | --- | --- | --- | --- | --- | --- |
| Model | Sampling Rate | Recall | IOU | Precision | PE Length (sec) | Predicted Length (sec) |
| GPT-4o | 1 | 0.998 [0.994, 1.000] | 0.784 [0.765, 0.803] | 0.792 [0.774, 0.811] | 126 [121, 132] | 175 [165, 187] |
| GPT-4o | 2 | 0.991 [0.985, 0.996] | 0.809 [0.794, 0.824] | 0.822 [0.808, 0.838] | 126 [121, 132] | 159 [150, 167] |
| GPT-4o | 3 | 0.979 [0.970, 0.986] | 0.796 [0.778, 0.813] | 0.817 [0.802, 0.833] | 126 [121, 132] | 157 [149, 166] |
| GPT-4o-mini | 1 | 0.945 [0.933, 0.956] | 0.835 [0.820, 0.849] | 0.878 [0.863, 0.894] | 126 [121, 132] | 141 [133, 149] |
| GPT-4o-mini | 2 | 0.918 [0.903, 0.931] | 0.822 [0.805, 0.838] | 0.885 [0.871, 0.900] | 126 [121, 132] | 134 [127, 141] |
| GPT-4o-mini | 3 | 0.886 [0.868, 0.903] | 0.791 [0.771, 0.809] | 0.880 [0.864, 0.894] | 126 [121, 132] | 130 [123, 137] |
| Gemini-2.0-Flash | 1 | **0.999 [0.999, 1.000]** | 0.631 [0.607, 0.656] | 0.639 [0.616, 0.667] | 126 [121, 132] | 262 [245, 277] |
| Gemini-2.0-Flash | 2 | 0.996 [0.990, 0.999] | 0.719 [0.699, 0.740] | 0.728 [0.708, 0.750] | 126 [121, 132] | 202 [188, 214] |
| Gemini-2.0-Flash | 3 | 0.993 [0.986, 0.998] | 0.722 [0.701, 0.743] | 0.731 [0.711, 0.751] | 126 [121, 132] | 197 [185, 210] |
| Gemma-3 | 1 | 0.995 [0.990, 0.998] | 0.531 [0.506, 0.558] | 0.541 [0.514, 0.571] | 126 [121, 132] | 332 [309, 353] |
| Gemma-3 | 2 | 0.993 [0.987, 0.997] | 0.621 [0.596, 0.647] | 0.631 [0.605, 0.658] | 126 [121, 132] | 270 [250, 288] |
| Gemma-3 | 3 | 0.989 [0.983, 0.994] | 0.634 [0.611, 0.656] | 0.647 [0.623, 0.673] | 126 [121, 132] | 254 [235, 270] |
| Qwen-2.5VL-72b | 1 | 0.919 [0.903, 0.934] | 0.765 [0.741, 0.784] | 0.827 [0.806, 0.851] | 126 [121, 132] | 166 [155, 178] |
| Qwen-2.5VL-72b | 2 | 0.879 [0.857, 0.899] | 0.752 [0.729, 0.774] | 0.846 [0.826, 0.867] | 126 [121, 132] | 152 [140, 162] |
| Qwen-2.5VL-72b | 3 | 0.847 [0.823, 0.869] | 0.736 [0.714, 0.757] | 0.854 [0.837, 0.875] | 126 [121, 132] | 139 [130, 148] |
| Qwen-2.5VL-7b | 1 | 0.929 [0.914, 0.943] | 0.425 [0.401, 0.450] | 0.462 [0.436, 0.493] | 126 [121, 132] | 382 [361, 400] |
| Qwen-2.5VL-7b | 2 | 0.881 [0.861, 0.902] | 0.470 [0.445, 0.493] | 0.531 [0.503, 0.559] | 126 [121, 132] | 319 [299, 338] |
| Qwen-2.5VL-7b | 3 | 0.860 [0.837, 0.884] | 0.475 [0.449, 0.501] | 0.547 [0.520, 0.580] | 126 [121, 132] | 299 [278, 319] |

Of all models evaluated, GPT-4o, Gemini-2.0-Flash, and Gemma-3 achieved the highest recall (0.998, 0.999, and 0.995, respectively) at a sampling rate of 1. We selected GPT-4o as our optimal model due to its superior combination of high recall with the best IOU (0.784), precision (0.792), and lowest predicted length (175 seconds) among high-recall performers.

We observed that increasing sampling rates generally reduced recall while improving IOU and precision. This inverse relationship between sampling rate and recall demonstrates how sampling frequency affects detection performance, with more frequent sampling capturing more examination events but potentially introducing more false positives.

### Classification Distribution Analysis

Figure 4 illustrates classification distributions across different time segments, revealing distinct patterns among models. In the three primary states analysis, all models showed relatively consistent classification patterns, with GPT-4o-mini and Qwen-2.5VL-72b notably classifying more “No Doctor” segments as “Preparation” compared to other models.

**Figure 4.**
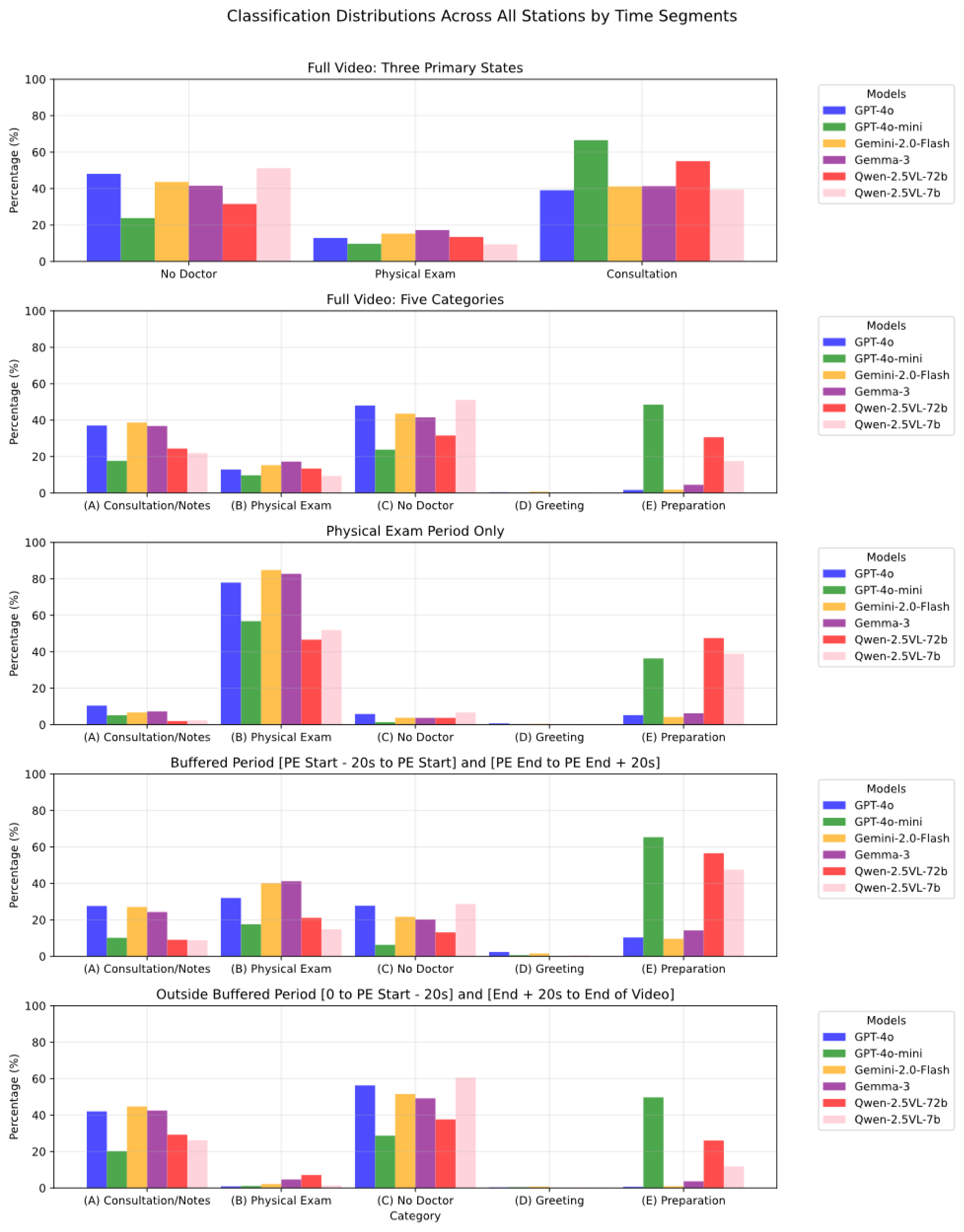
Classification Distribution Percentages by Station. (A) Taking Notes or Consulting, (B) Physical Exam, (C) No Doctor, (D) Greeting, and (E) Preparation.

During physical examination periods, all models correctly identified a high percentage of frames as “Physical Exam.” The buffer period analysis (±20 seconds) showed a gradual transition between states, while classifications outside the buffered period correctly showed minimal “Physical Exam” identifications, confirming the models’ spatial accuracy.

### Station-Specific Performance at 1 FPS

We examined performance variations across the five different OSCE station types to evaluate our approach’s adaptability to different clinical scenarios. Figure 5 demonstrates that GPT-4o achieved high recall performance across all stations, with most cases achieving near-perfect recall (100%).

**Figure 5.**
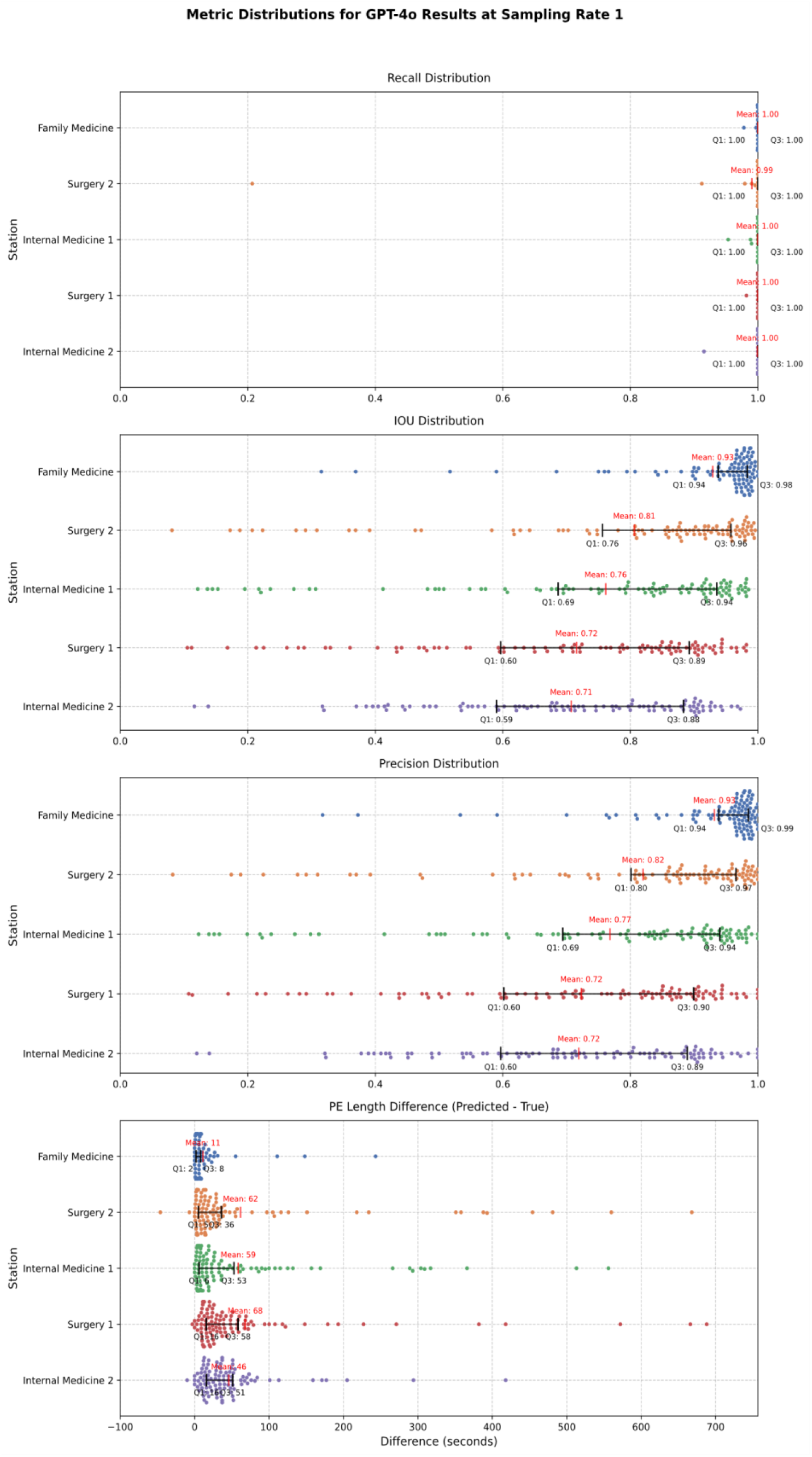
Recall, IOU, Physical Exam Length Difference Distributions with IQR by Station for GPT-4o

The Family Medicine station exhibited the highest IOU (mean: 0.93), likely because this station did not require medical students to place patients in a supine position for assessment, which reduced false positive physical examination classifications. Surgery 1 and Internal Medicine 2 stations showed the lowest IOU means (0.72 and 0.71, respectively), though they still maintained strong performance.

The PE Length Difference analysis in the bottom panel of Figure 5 indicates that GPT-4o generally predicted slightly longer examination periods than the ground truth across all stations, with mean differences ranging from 11 seconds (Family Medicine) to 68 seconds (Surgery 1). These extended predictions ensure a comprehensive capture of examination activities while maintaining manageable review segments.

### Classification Error Analysis

We identified two primary types of error cases that affected our model’s performance: false negatives that reduced recall and false positives that reduced precision and IOU. Table 5 categorizes these errors into five distinct failure modes with representative examples:

**Table 5.**
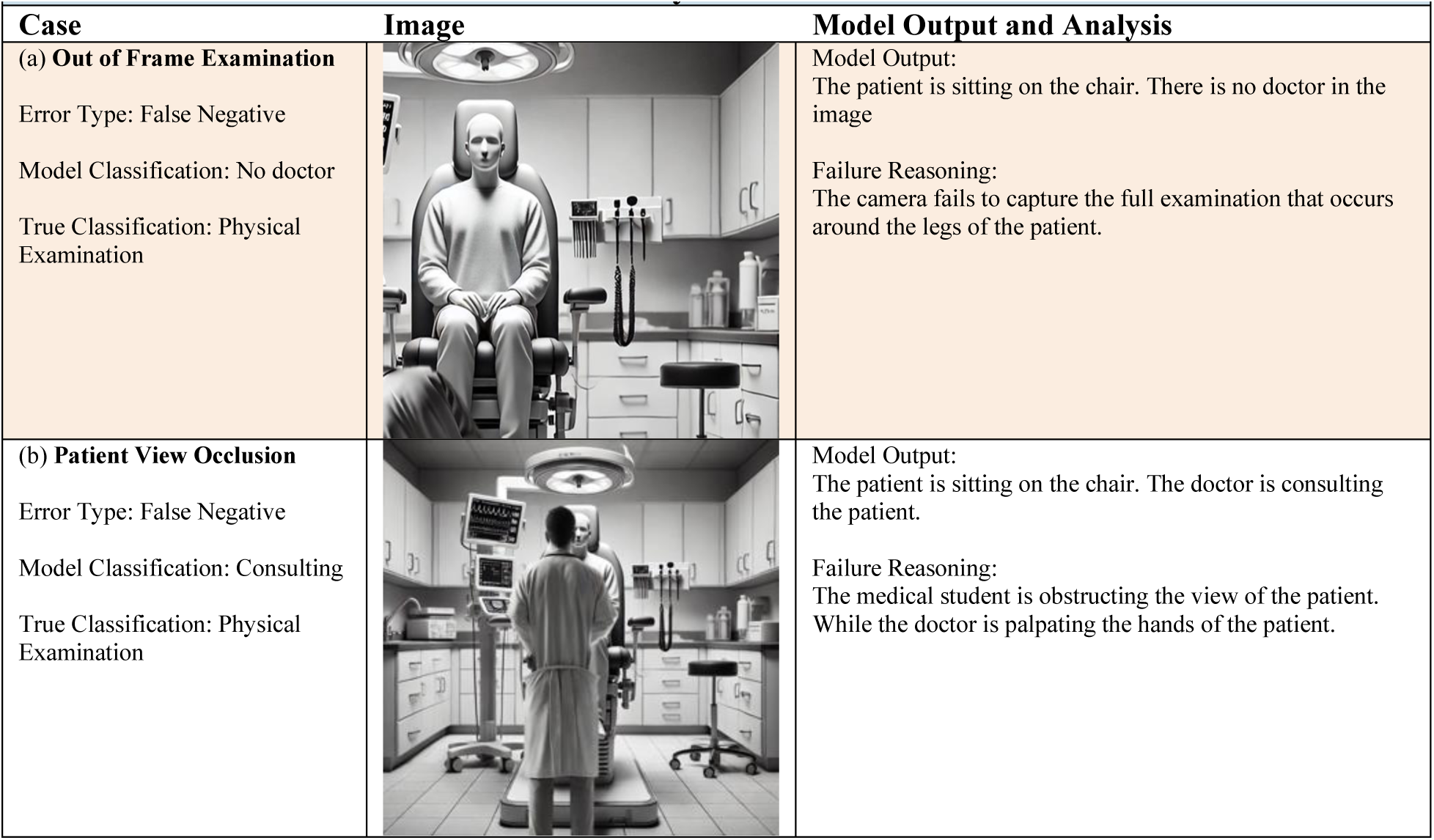
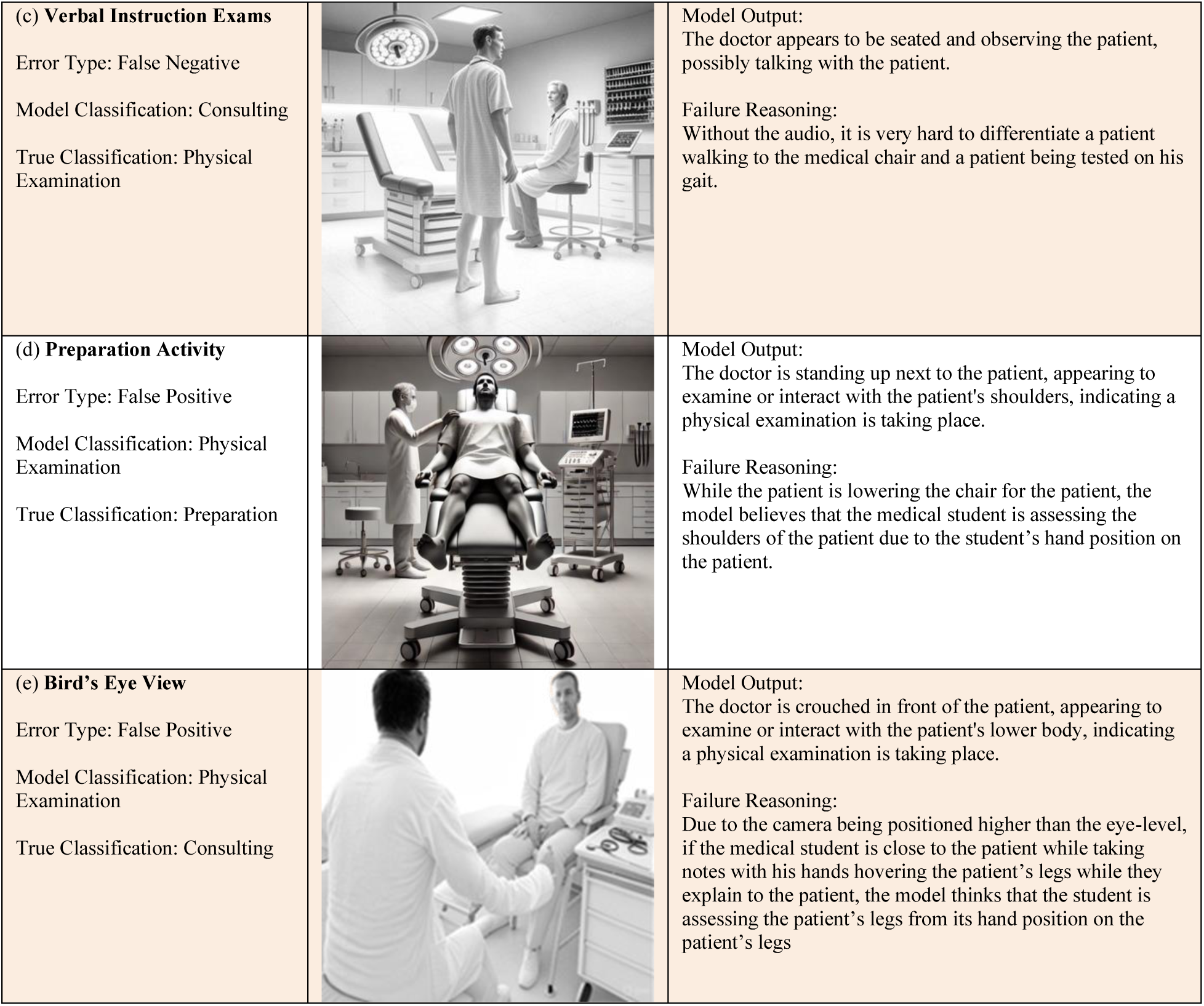
Classification Failure Mode Analysis.

| Table 5. Classification Failure Mode Analysis |  |  |
| --- | --- | --- |
| Case | Image | Model Output and Analysis |
| (a) **Out of Frame Examination**<br>Error Type: False Negative<br>Model Classification: No doctor<br>True Classification: Physical Examination |  | Model Output:<br>The patient is sitting on the chair. There is no doctor in the image<br>Failure Reasoning:<br>The camera fails to capture the full examination that occurs around the legs of the patient. |
| (b) **Patient View Occlusion**<br>Error Type: False Negative<br>Model Classification: Consulting<br>True Classification: Physical Examination |  | Model Output:<br>The patient is sitting on the chair. The doctor is consulting the patient.<br>Failure Reasoning:<br>The medical student is obstructing the view of the patient. While the doctor is palpating the hands of the patient. |
| (c) **Verbal Instruction Exams**<br>Error Type: False Negative<br>Model Classification: Consulting<br>True Classification: Physical Examination |  | Model Output:<br>The doctor appears to be seated and observing the patient, possibly talking with the patient.<br>Failure Reasoning:<br>Without the audio, it is very hard to differentiate a patient walking to the medical chair and a patient being tested on his gait. |
| (d) **Preparation Activity**<br>Error Type: False Positive<br>Model Classification: Physical Examination<br>True Classification: Preparation |  | Model Output:<br>The doctor is standing up next to the patient, appearing to examine or interact with the patient's shoulders, indicating a physical examination is taking place.<br>Failure Reasoning:<br>While the patient is lowering the chair for the patient, the model believes that the medical student is assessing the shoulders of the patient due to the student's hand position on the patient. |
| (e) **Bird's Eye View**<br>Error Type: False Positive<br>Model Classification: Physical Examination<br>True Classification: Consulting |  | Model Output:<br>The doctor is crouched in front of the patient, appearing to examine or interact with the patient's lower body, indicating a physical examination is taking place.<br>Failure Reasoning:<br>Due to the camera being positioned higher than the eye-level, if the medical student is close to the patient while taking notes with his hands hovering the patient's legs while they explain to the patient, the model thinks that the student is assessing the patient's legs from its hand position on the patient's legs |

All images used in the error analysis (Table 5) were generated by Grok3 to protect patient and student privacy while accurately representing the classification challenges observed in the actual OSCE videos.

These failure modes provide valuable insights for future system optimizations, particularly regarding camera placement and the potential integration of additional contextual information.

## Discussion

This study demonstrates that integrating MM-LMMs with temporal modeling provides an effective solution for automatically segmenting physical examination periods within OSCE videos. Our approach achieved 99.8% recall while reducing the video content requiring review by 81%, offering substantial benefits for both manual and automated assessment of medical student clinical skills. Below, we discuss the implications, limitations, and future directions of this work.

### Significance for Medical Education Assessment

The high recall rate (99.8%) achieved by our GPT-4o implementation represents a critical advancement for clinical skills assessment. In medical education, missing even brief examination components can lead to incomplete evaluations, potentially overlooking critical errors or skillful techniques that determine pass/fail outcomes. Our approach ensures comprehensive capture of examination activities while substantially reducing the review burden.

By reducing a 15-minute (900-second) video to an average of 175 seconds of relevant content, our method creates significant efficiency gains for educational institutions. For programs conducting OSCEs with hundreds of students across multiple stations, this could translate to significant time savings, potentially thousands of hours saved annually. Beyond time savings, this approach enhances assessment quality by allowing evaluators to focus their attention on the most pertinent segments of student performance.

The station-specific performance analysis revealed important insights about contextual factors affecting detection accuracy. Family Medicine stations achieved the highest precision (93.2%) and IOU (93.4%), likely due to their seated examination format with minimal patient repositioning. In contrast, Surgery stations showed more variability in precision (72.3%), reflecting the challenges posed by frequent patient repositioning and varying examination techniques. These findings highlight how physical examination contexts influence detection performance and suggest potential optimization strategies for different clinical scenarios.

### Practical Implementation and Limitations

Implementation in educational settings should begin with optimizing camera positioning. Our error analysis identified several failure modes directly attributable to suboptimal camera angles, including out-of-frame examinations and patient view occlusion. Based on these findings, we recommend positioning cameras at eye level with a side view that captures both the medical student and patient simultaneously.

Institutions must also consider data privacy regulations when implementing AI-based video analysis systems. Our research used HIPAA- and FERPA-compliant APIs for proprietary models, while open-source alternatives were hosted on secure institutional infrastructure. These approaches demonstrate viable pathways for maintaining compliance while leveraging advanced AI capabilities in educational settings.

Several limitations warrant consideration. Dynamic assessment activities, such as gait examinations, pose challenges for frame-based classification since a single frame cannot reliably distinguish between a patient walking as part of an examination versus walking to the examination bed. For such specialized examinations, supplementing visual analysis with transcript-based temporal markers would likely improve classification accuracy.

Additionally, our approach does not currently differentiate between specific examination techniques (e.g., palpation, auscultation, percussion). While the current implementation successfully identifies general physical examination periods, a more granular classification would further enhance feedback specificity for competency-based education.

### Future Directions

Several promising directions emerge for extending this work. First, fine-tuning open-source MM-LLMs on domain-specific datasets could improve boundary detection by incorporating prior knowledge of examination protocols. This approach would allow institutions without access to HIPAA and FERPA-compliant APIs to implement this method locally.

Second, multi-modal fusion approaches that combine visual analysis with audio transcripts could address the limitations identified for dynamic examinations. Integrating transcript markers for examination commands (e.g., “Please walk across the room”) with visual cues could improve detection accuracy for ambiguous activities.

Third, expanding the classification granularity to identify specific examination techniques would enable more detailed feedback on clinical skills. This could involve developing hierarchical models that first identify general examination periods before classifying specific techniques within those segments.

Finally, this framework could extend beyond OSCEs to other time-intensive healthcare education scenarios, such as surgical skills assessment, emergency simulation training, or telemedicine encounters. The principles of combining zero-shot visual classification with temporal modeling could generalize to these domains, offering similar efficiency gains for educational assessment.

## Conclusion

Our study demonstrates that integrating MM-LLMs with Viterbi decoding creates an effective solution for automatically segmenting physical examination periods within OSCE videos. By achieving 99.8% recall while reducing video content requiring review by 81%, our approach offers substantial efficiency gains for both manual and automated assessment of medical student clinical skills. This high recall rate is particularly critical in educational contexts, where missing even brief examination components could lead to incomplete evaluations of student competence.

Beyond immediate efficiency improvements, this work establishes a framework for a more sophisticated automated assessment of clinical skills. As medical education increasingly adopts competency-based frameworks demanding more frequent, detailed assessments, such technological innovations will be essential for balancing educational quality with institutional resources. By enabling a more targeted review of student performance, our approach not only reduces faculty workload but potentially enhances assessment quality by allowing evaluators to focus their expertise on the most relevant segments of student performance. This technical advancement represents an important step toward comprehensive, scalable, and objective assessment of clinical competencies in medical education.

## Data Availability

Data will not be available

## Notes

### Competing Interest Statement

The authors have declared no competing interest.

### Funding Statement

Azure compute credits were provided to Dr. Jamieson by Microsoft as part of the Accelerating Foundation Models Research initiative.

### Author Declarations

IRB of UT Southwestern Medical Center gave ethical approval for this work

## References

1. Hodges, B. (2003). OSCE! Variations on a theme by Harden. Medical Education, 37(12), 1134–1140. 10.1111/j.1365-2923.2003.01717.x

2. Harden, R. M., Stevenson, M., Downie, W. W., & Wilson, G. M. (1975). Assessment of clinical competence using objective structured examination. British Medical Journal, 1(5955), 447–451. 10.1136/bmj.1.5955.447

3. Regehr, G., MacRae, H., Reznick, R. K., & Szalay, D. (1998). Comparing the psychometric properties of checklists and global rating scales for assessing performance on an OSCE-format examination. Academic Medicine: Journal of the Association of American Medical Colleges, 73(9), 993–997. 10.1097/00001888-199809000-00020

4. Jamieson, A. R., Holcomb, M. J., Dalton, T. O., Campbell, K. K., Vedovato, S., Shakur, A. H., Kang, S., Hein, D., Lawson, J., Danuser, G., & Scott, D. J. (2024). Rubrics to Prompts: Assessing Medical Student Post-Encounter Notes with AI. NEJM AI, 1(12). 10.1056/AIcs2400631

5. Shakur, A. H., Holcomb, M. J., Hein, D., Kang, S., Dalton, T. O., Campbell, K. K., Scott, D. J., & Jamieson, A. R. (2024). Large Language Models for Medical OSCE Assessment: A Novel Approach to Transcript Analysis. arXiv:2410.12858. 10.48550/arXiv.2410.12858

6. Vedovato, S., Kang, S., Holcomb, M. J., Campbell, K. K., Scott, D. J., Dalton, T. O., Danuser, G., & Jamieson, A. R. (2024). Towards Better Debriefing Through Context-Aware Video Segmentation in Standardized Patient Encounter Ear Exams. 2024 IEEE First International Conference on Artificial Intelligence for Medicine, Health and Care (AIMHC), 162–165. 10.1109/AIMHC59811.2024.00036

7. Khan, K. Z., Ramachandran, S., Gaunt, K., & Pushkar, P. (2013). The Objective Structured Clinical Examination (OSCE): AMEE Guide No. 81. Part I: an historical and theoretical perspective. Medical Teacher, 35(9), e1437–1446. 10.3109/0142159X.2013.818634

8. Epstein, R. M. (2007). Assessment in medical education. The New England Journal of Medicine, 356(4), 387–396. 10.1056/NEJMra054784

9. Qian, D., Ji, H., Hong, X., Yan, B., Chen, Z., Zhu, X., Tian, J., Duan, S., & Zhou, X. (2024). VideoStreaming: Understanding Videos of Arbitrary Length with Vision-Language Models. arXiv preprint arXiv:2402.12195.

10. Ren, Z., Zheng, C., Wang, Y., Li, S., Lin, Y., Liu, Z., Gan, C., & Yu, Z. (2024). Memory-Augmented Large Multimodal Model for Long-Term Video Understanding. arXiv preprint arXiv:2401.10965.

11. Zou, H., Luo, T., Xie, G., Zhang, V., Lv, F., Wang, G., Chen, J., Wang, Z., Zhang, H., & Zhang, H. (2024). From Seconds to Hours: Reviewing MultiModal Large Language Models on Comprehensive Long Video Understanding. arXiv preprint arXiv:2409.18938.

12. Karpathy, A., Toderici, G., Shetty, S., Leung, T., Sukthankar, R., & Fei-Fei, L. (2014). Large-Scale Video Classification with Convolutional Neural Networks. In 2014 IEEE Conference on Computer Vision and Pattern Recognition, 1725–32. 10.1109/CVPR.2014.223.

13. Patil, P., Pawar, V., Pawar, Y., & Pisal, S. (2021). Video Content Classification Using Deep Learning. arXiv preprint arXiv:2111.13813. 10.48550/arXiv.2111.13813.

14. Ding, G., Sener, F., & Yao, A. (2023). Temporal Action Segmentation: An Analysis of Modern Techniques. arXiv preprint arXiv:2210.10352. 10.48550/arXiv.2210.10352.

15. YOLOv11: An Overview of the Key Architectural Enhancements. (n.d.). Retrieved March 1, 2025, from https://arxiv.org/html/2410.17725v1

16. Newell, A., Yang, K., & Deng, J. (2016). Stacked Hourglass Networks for Human Pose Estimation. arXiv preprint arXiv:1603.06937. 10.48550/arXiv.1603.06937.

17. Moreno-Pino, F., Sükei, E., Olmos, P. M., & Artés-Rodríguez, A. (2022). PyHHMM: A Python Library for Heterogeneous Hidden Markov Models. arXiv preprint arXiv:2201.06968. 10.48550/arXiv.2201.06968.

