## Appendix for "Automatic Physical Examination Segmentation within Objective Structured Clinical Examination Videos"

A.

| Appendix A. Performance Metrics with Buffer with 95% Confidence Intervals | | | | | | | |
|---|---|---|---|---|---|---|---|
| **Model** | **Sampling Rate** | **Buffer** | **Recall** | **IOU** | **Precision** | **PE Length (sec)** | **Predicted Length (sec)** |
| GPT-4o | 1 | 0s | 0.998 [0.994, 1.000] | 0.784 [0.765, 0.803] | 0.792 [0.774, 0.811] | 126 [121, 132] | 175 [165, 187] |
| GPT-4o | 1 | 15s | 0.999 [0.998, 1.000] | 0.654 [0.637, 0.670] | 0.664 [0.648, 0.680] | 126 [121, 132] | 202 [192, 214] |
| GPT-4o | 1 | 30s | 1.000 [1.000, 1.000] | 0.559 [0.545, 0.573] | 0.571 [0.556, 0.586] | 126 [121, 132] | 232 [221, 243] |
| GPT-4o | 2 | 0s | 0.991 [0.985, 0.996] | 0.809 [0.794, 0.824] | 0.822 [0.808, 0.838] | 126 [121, 132] | 159 [150, 167] |
| GPT-4o | 2 | 15s | 0.996 [0.993, 0.999] | 0.684 [0.670, 0.697] | 0.696 [0.682, 0.711] | 126 [121, 132] | 184 [176, 193] |
| GPT-4o | 2 | 30s | 0.999 [0.997, 1.000] | 0.584 [0.571, 0.598] | 0.597 [0.584, 0.610] | 126 [121, 132] | 213 [205, 222] |
| GPT-4o | 3 | 0s | 0.979 [0.970, 0.986] | 0.796 [0.778, 0.813] | 0.817 [0.802, 0.833] | 126 [121, 132] | 157 [149, 166] |
| GPT-4o | 3 | 15s | 0.987 [0.980, 0.993] | 0.688 [0.672, 0.703] | 0.703 [0.689, 0.719] | 126 [121, 132] | 180 [172, 189] |
| GPT-4o | 3 | 30s | 0.993 [0.987, 0.997] | 0.591 [0.576, 0.605] | 0.604 [0.590, 0.619] | 126 [121, 132] | 210 [201, 219] |
| GPT-4o-mini | 1 | 0s | 0.945 [0.933, 0.956] | 0.835 [0.820, 0.849] | 0.878 [0.863, 0.894] | 126 [121, 132] | 141 [133, 149] |
| GPT-4o-mini | 1 | 15s | 0.974 [0.963, 0.982] | 0.719 [0.704, 0.733] | 0.737 [0.721, 0.753] | 126 [121, 132] | 168 [161, 176] |
| GPT-4o-mini | 1 | 30s | 0.984 [0.975, 0.991] | 0.616 [0.603, 0.630] | 0.628 [0.613, 0.643] | 126 [121, 132] | 198 [190, 205] |
| GPT-4o-mini | 2 | 0s | 0.918 [0.903, 0.931] | 0.822 [0.805, 0.838] | 0.885 [0.871, 0.900] | 126 [121, 132] | 134 [127, 141] |
| GPT-4o-mini | 2 | 15s | 0.955 [0.943, 0.965] | 0.728 [0.714, 0.742] | 0.757 [0.741, 0.772] | 126 [121, 132] | 159 [152, 167] |
| GPT-4o-mini | 2 | 30s | 0.974 [0.963, 0.982] | 0.632 [0.618, 0.644] | 0.647 [0.631, 0.662] | 126 [121, 132] | 188 [181, 196] |

| | | | | | | | |
|---|---|---|---|---|---|---|---|
| **GPT-4o-mini** | **3** | **0s** | **0.886 [0.868, 0.903]** | **0.791 [0.771, 0.809]** | **0.880 [0.865, 0.894]** | **126 [121, 132]** | **130 [123, 137]** |
| **GPT-4o-mini** | **3** | **15s** | **0.931 [0.916, 0.945]** | **0.723 [0.707, 0.737]** | **0.767 [0.752, 0.782]** | **126 [121, 132]** | **153 [146, 161]** |
| **GPT-4o-mini** | **3** | **30s** | **0.959 [0.947, 0.970]** | **0.636 [0.621, 0.650]** | **0.658 [0.643, 0.673]** | **126 [121, 132]** | **183 [175, 190]** |
| **Gemini-2.0-Flash** | **1** | **0s** | **0.999 [0.999, 1.000]** | **0.631 [0.607, 0.656]** | **0.639 [0.612, 0.667]** | **126 [121, 132]** | **262 [245, 277]** |
| **Gemini-2.0-Flash** | **1** | **15s** | **1.000 [1.000, 1.000]** | **0.536 [0.516, 0.557]** | **0.546 [0.527, 0.569]** | **126 [121, 132]** | **289 [273, 305]** |
| **Gemini-2.0-Flash** | **1** | **30s** | **1.000 [1.000, 1.000]** | **0.465 [0.449, 0.483]** | **0.477 [0.460, 0.496]** | **126 [121, 132]** | **318 [302, 334]** |
| **Gemini-2.0-Flash** | **2** | **0s** | **0.996 [0.990, 0.999]** | **0.719 [0.699, 0.740]** | **0.728 [0.706, 0.750]** | **126 [121, 132]** | **202 [188, 214]** |
| **Gemini-2.0-Flash** | **2** | **15s** | **0.998 [0.993, 1.000]** | **0.613 [0.597, 0.632]** | **0.624 [0.606, 0.640]** | **126 [121, 132]** | **227 [214, 240]** |
| **Gemini-2.0-Flash** | **2** | **30s** | **0.998 [0.995, 1.000]** | **0.527 [0.513, 0.543]** | **0.540 [0.524, 0.557]** | **126 [121, 132]** | **257 [243, 269]** |
| **Gemini-2.0-Flash** | **3** | **0s** | **0.993 [0.986, 0.998]** | **0.722 [0.701, 0.743]** | **0.731 [0.711, 0.751]** | **126 [121, 132]** | **197 [185, 210]** |
| **Gemini-2.0-Flash** | **3** | **15s** | **0.995 [0.989, 0.999]** | **0.623 [0.605, 0.642]** | **0.633 [0.616, 0.653]** | **126 [121, 132]** | **223 [209, 233]** |
| **Gemini-2.0-Flash** | **3** | **30s** | **0.996 [0.990, 1.000]** | **0.535 [0.520, 0.551]** | **0.547 [0.531, 0.563]** | **126 [121, 132]** | **250 [237, 263]** |
| **Gemma-3** | **1** | **0s** | **0.995 [0.990, 0.998]** | **0.531 [0.506, 0.558]** | **0.541 [0.514, 0.571]** | **126 [121, 132]** | **332 [309, 353]** |
| **Gemma-3** | **1** | **15s** | **0.997 [0.993, 1.000]** | **0.460 [0.439, 0.482]** | **0.470 [0.449, 0.495]** | **126 [121, 132]** | **359 [336, 380]** |
| **Gemma-3** | **1** | **30s** | **0.998 [0.995, 1.000]** | **0.405 [0.387, 0.423]** | **0.416 [0.399, 0.437]** | **126 [121, 132]** | **387 [365, 409]** |
| **Gemma-3** | **2** | **0s** | **0.993 [0.987, 0.997]** | **0.621 [0.596, 0.647]** | **0.631 [0.605, 0.658]** | **126 [121, 132]** | **270 [250, 288]** |
| **Gemma-3** | **2** | **15s** | **0.996 [0.991, 0.999]** | **0.538 [0.516, 0.559]** | **0.548 [0.526, 0.569]** | **126 [121, 132]** | **295 [275, 314]** |
| **Gemma-3** | **2** | **30s** | **0.997 [0.992, 1.000]** | **0.468 [0.449, 0.487]** | **0.479 [0.460, 0.497]** | **126 [121, 132]** | **324 [304, 342]** |
| **Gemma-3** | **3** | **0s** | **0.989 [0.983, 0.994]** | **0.634 [0.611, 0.656]** | **0.647 [0.623, 0.673]** | **126 [121, 132]** | **254 [235, 270]** |
| **Gemma-3** | **3** | **15s** | **0.994 [0.989, 0.998]** | **0.554 [0.534, 0.574]** | **0.567 [0.546, 0.588]** | **126 [121, 132]** | **277 [258, 293]** |

| | | | | | | | |
|---|---|---|---|---|---|---|---|
| Gemma-3 | 3 | 30s | 0.997 [0.993, 0.999] | 0.481 [0.464, 0.499] | 0.494 [0.476, 0.512] | 126 [121, 132] | 306 [287, 322] |
| Qwen-2.5VL-72b | 1 | 0s | 0.919 [0.903, 0.934] | 0.765 [0.741, 0.785] | 0.827 [0.806, 0.851] | 126 [121, 132] | 166 [155, 178] |
| Qwen-2.5VL-72b | 1 | 15s | 0.956 [0.943, 0.967] | 0.673 [0.654, 0.691] | 0.701 [0.682, 0.722] | 126 [121, 132] | 193 [182, 205] |
| Qwen-2.5VL-72b | 1 | 30s | 0.974 [0.963, 0.983] | 0.586 [0.568, 0.603] | 0.604 [0.585, 0.622] | 126 [121, 132] | 222 [211, 234] |
| Qwen-2.5VL-72b | 2 | 0s | 0.879 [0.857, 0.899] | 0.752 [0.729, 0.774] | 0.846 [0.828, 0.867] | 126 [121, 132] | 152 [140, 162] |
| Qwen-2.5VL-72b | 2 | 15s | 0.925 [0.906, 0.940] | 0.681 [0.663, 0.698] | 0.729 [0.712, 0.748] | 126 [121, 132] | 177 [165, 187] |
| Qwen-2.5VL-72b | 2 | 30s | 0.953 [0.939, 0.965] | 0.602 [0.586, 0.617] | 0.629 [0.612, 0.647] | 126 [121, 132] | 206 [194, 216] |
| Qwen-2.5VL-72b | 3 | 0s | 0.847 [0.823, 0.869] | 0.736 [0.714, 0.757] | 0.854 [0.837, 0.875] | 126 [121, 132] | 139 [130, 148] |
| Qwen-2.5VL-72b | 3 | 15s | 0.895 [0.874, 0.912] | 0.679 [0.661, 0.697] | 0.747 [0.729, 0.765] | 126 [121, 132] | 162 [153, 171] |
| Qwen-2.5VL-72b | 3 | 30s | 0.934 [0.917, 0.947] | 0.610 [0.595, 0.626] | 0.647 [0.629, 0.665] | 126 [121, 132] | 191 [182, 200] |
| Qwen-2.5VL-7b | 1 | 0s | 0.929 [0.914, 0.943] | 0.425 [0.401, 0.450] | 0.462 [0.436, 0.493] | 126 [121, 132] | 382 [364, 400] |
| Qwen-2.5VL-7b | 1 | 15s | 0.958 [0.946, 0.969] | 0.394 [0.374, 0.416] | 0.416 [0.393, 0.442] | 126 [121, 132] | 407 [387, 426] |
| Qwen-2.5VL-7b | 1 | 30s | 0.973 [0.963, 0.982] | 0.361 [0.343, 0.381] | 0.376 [0.356, 0.398] | 126 [121, 132] | 434 [414, 452] |
| Qwen-2.5VL-7b | 2 | 0s | 0.881 [0.861, 0.902] | 0.470 [0.445, 0.493] | 0.531 [0.503, 0.559] | 126 [121, 132] | 319 [299, 338] |
| Qwen-2.5VL-7b | 2 | 15s | 0.918 [0.900, 0.935] | 0.444 [0.421, 0.465] | 0.479 [0.454, 0.503] | 126 [121, 132] | 343 [323, 362] |
| Qwen-2.5VL-7b | 2 | 30s | 0.946 [0.929, 0.960] | 0.410 [0.389, 0.429] | 0.431 [0.409, 0.453] | 126 [121, 132] | 370 [351, 389] |
| Qwen-2.5VL-7b | 3 | 0s | 0.860 [0.837, 0.884] | 0.475 [0.449, 0.504] | 0.547 [0.519, 0.577] | 126 [121, 132] | 299 [278, 320] |
| Qwen-2.5VL-7b | 3 | 15s | 0.897 [0.876, 0.918] | 0.452 [0.429, 0.477] | 0.499 [0.472, 0.527] | 126 [121, 132] | 321 [300, 341] |
| Qwen-2.5VL-7b | 3 | 30s | 0.924 [0.905, 0.942] | 0.419 [0.399, 0.441] | 0.449 [0.425, 0.474] | 126 [121, 132] | 348 [327, 367] |

B.

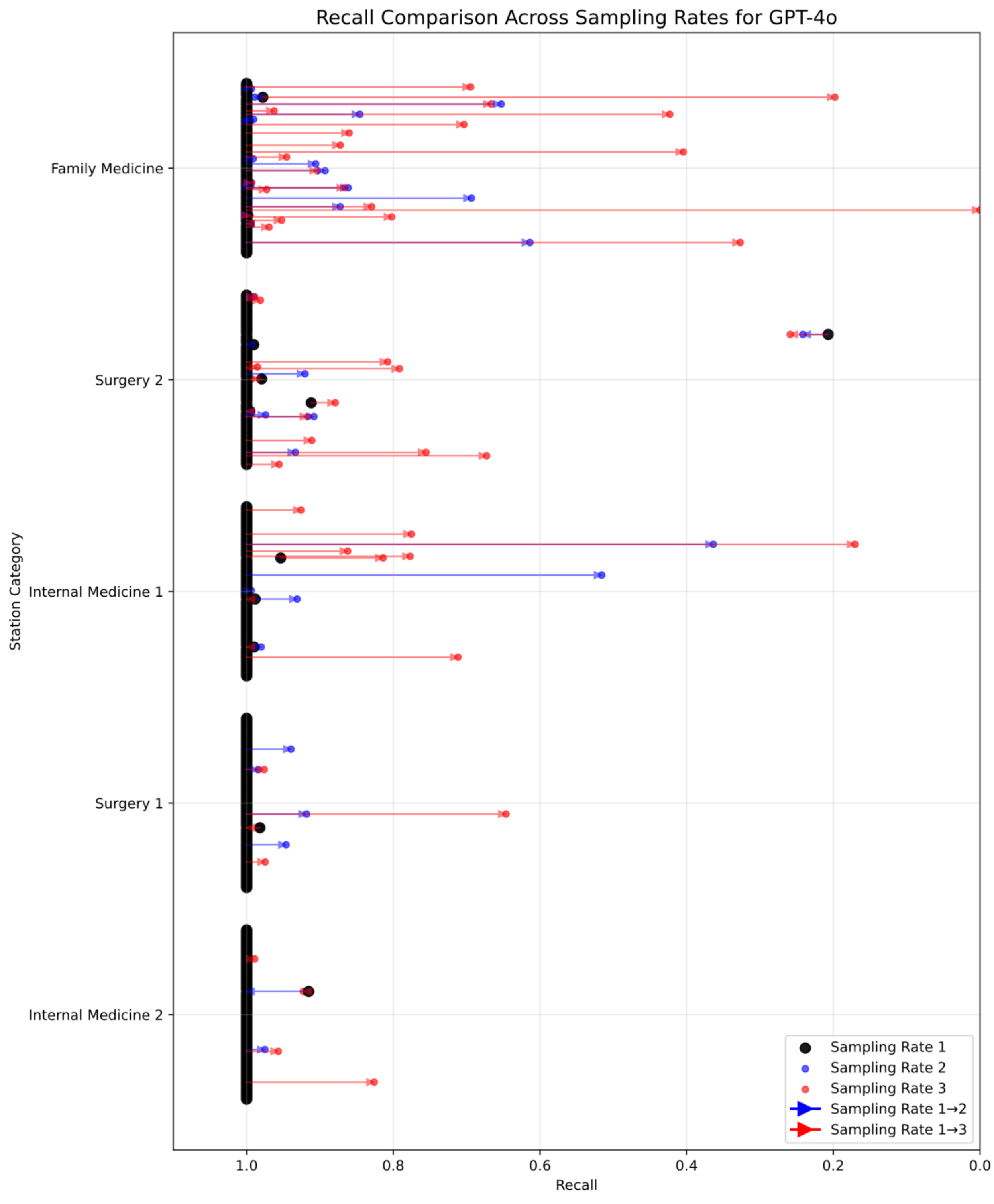

Recall Comparison Across Sampling Rates for GPT-4o
Family Medicine
Surgery 2
Internal Medicine 1
Surgery 1
Internal Medicine 2
Station Category
1.0
0.8
0.6
0.4
0.2
0.0
Recall
Sampling Rate 1
Sampling Rate 2
Sampling Rate 3
Sampling Rate 1→2
Sampling Rate 1→3


C.

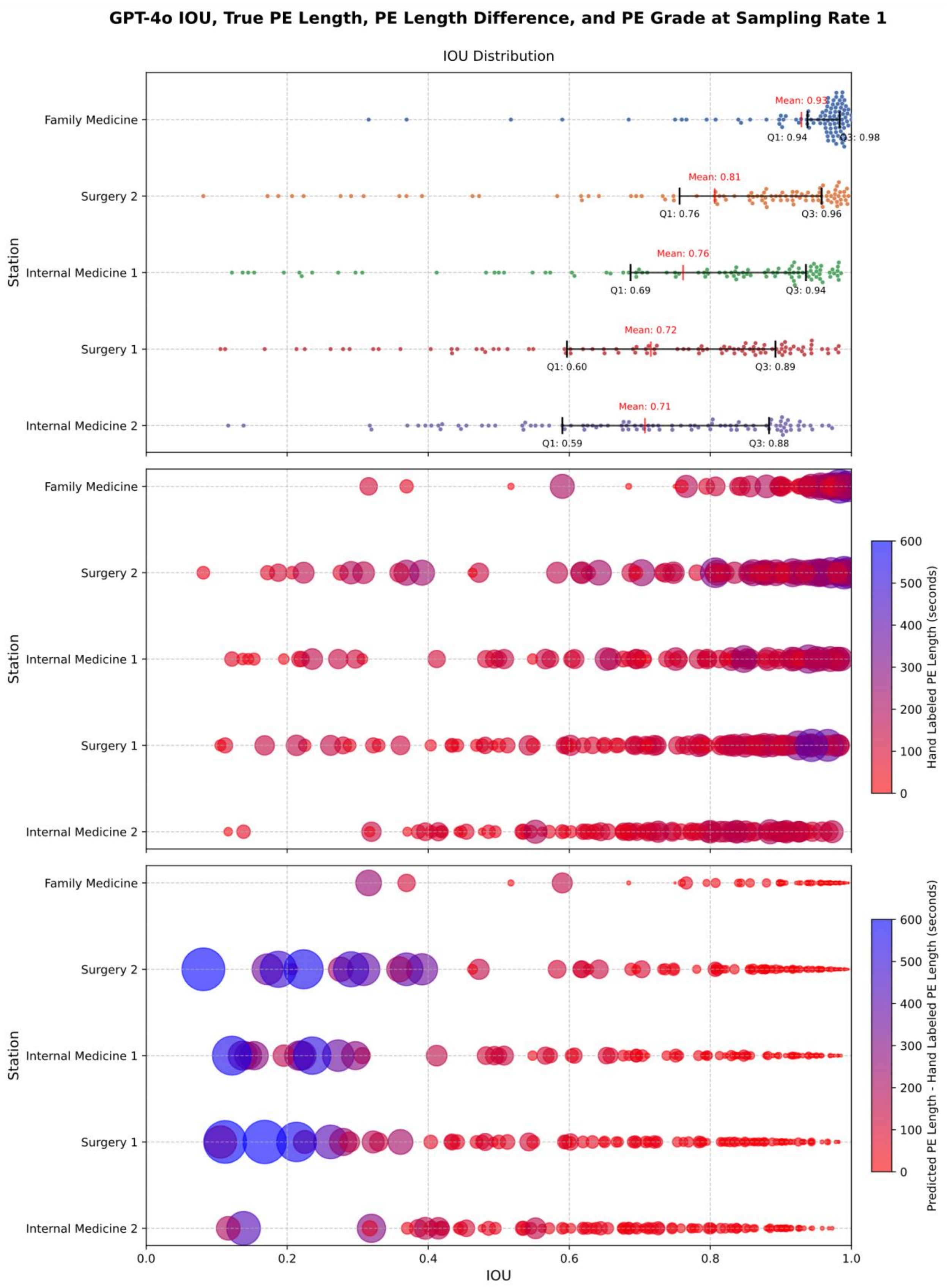
GPT-4o IOU, True PE Length, PE Length Difference, and PE Grade at Sampling Rate 1
IOU Distribution
Family Medicine
Surgery 2
Internal Medicine 1
Surgery 1
Internal Medicine 2
Station
Mean: 0.93
Q1: 0.94
Q3: 0.98
Mean: 0.81
Q1: 0.76
Q3: 0.96
Mean: 0.76
Q1: 0.69
Q3: 0.94
Mean: 0.72
Q1: 0.60
Q3: 0.89
Mean: 0.71
Q1: 0.59
Q3: 0.88
Hand Labeled PE Length (seconds)
Predicted PE Length - Hand Labeled PE Length (seconds)
0
100
200
300
400
500
600
0.0
0.2
0.4
0.6
0.8
1.0
IOU

D.

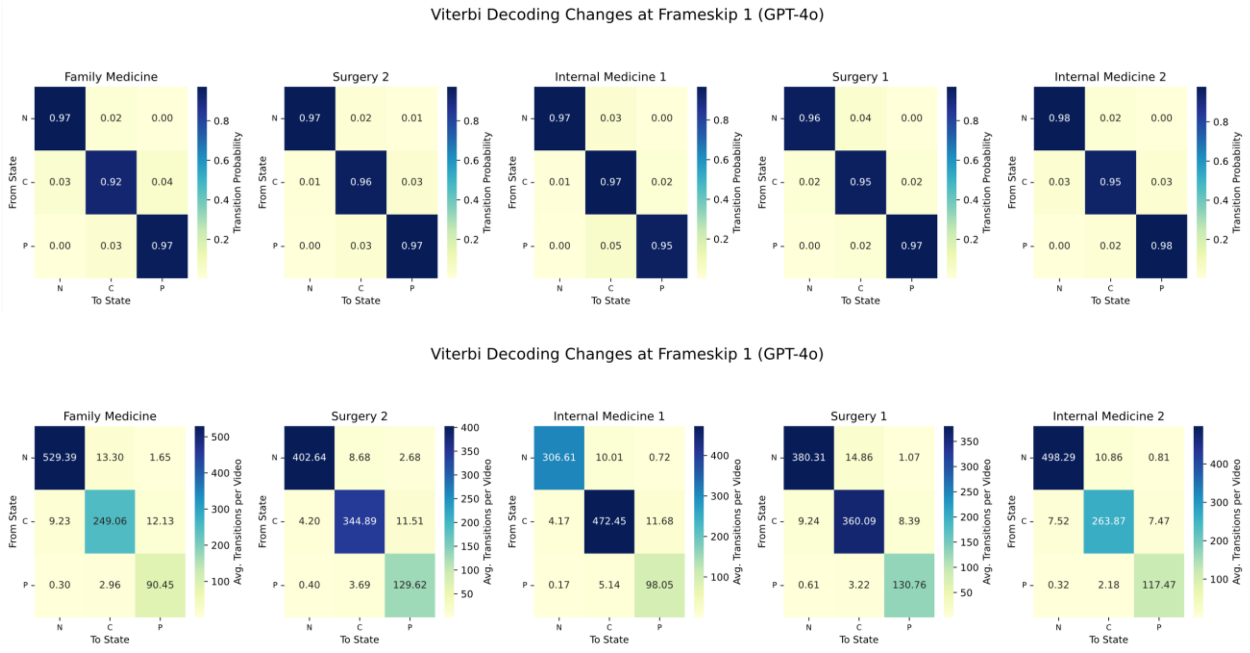


E.

**Table 4. Performance Metrics of GPT-4o Across Stations with 95% Confidence Intervals**

| Station | Sampling Rate | Recall | IOU | Precision | PE Length (sec) | Predicted Length (sec) |
|---|---|---|---|---|---|---|
| **Overall Average** | **1** | **0.998 [0.994, 1.000]** | **0.784 [0.765, 0.803]** | **0.792 [0.774, 0.811]** | **126 [121, 132]** | **175 [165, 187]** |
| Family Medicine | 1 | 1.000 [0.999, 1.000] | 0.929 [0.909, 0.950] | 0.932 [0.905, 0.951] | 140 [127, 154] | 151 [137, 165] |
| Surgery 2 | 1 | 0.991 [0.974, 1.000] | 0.806 [0.764, 0.853] | 0.820 [0.782, 0.864] | 161 [147, 173] | 222 [196, 246] |
| Internal Medicine 1 | 1 | 0.999 [0.998, 1.000] | 0.761 [0.721, 0.801] | 0.768 [0.727, 0.813] | 110 [101, 119] | 168 [147, 190] |
| Surgery 1 | 1 | 1.000 [0.999, 1.000] | 0.716 [0.673, 0.759] | 0.723 [0.680, 0.763] | 119 [108, 131] | 187 [166, 212] |
| Internal Medicine 2 | 1 | 0.999 [0.997, 1.000] | 0.707 [0.674, 0.745] | 0.719 [0.690, 0.750] | 101 [93, 107] | 147 [134, 160] |
